## Supplementary Information for "Microfluidic Affinity Profiling reveals a Broad Range of Target Affinities for Anti-SARS-CoV-2 Antibodies in Plasma of COVID-19 Survivors"

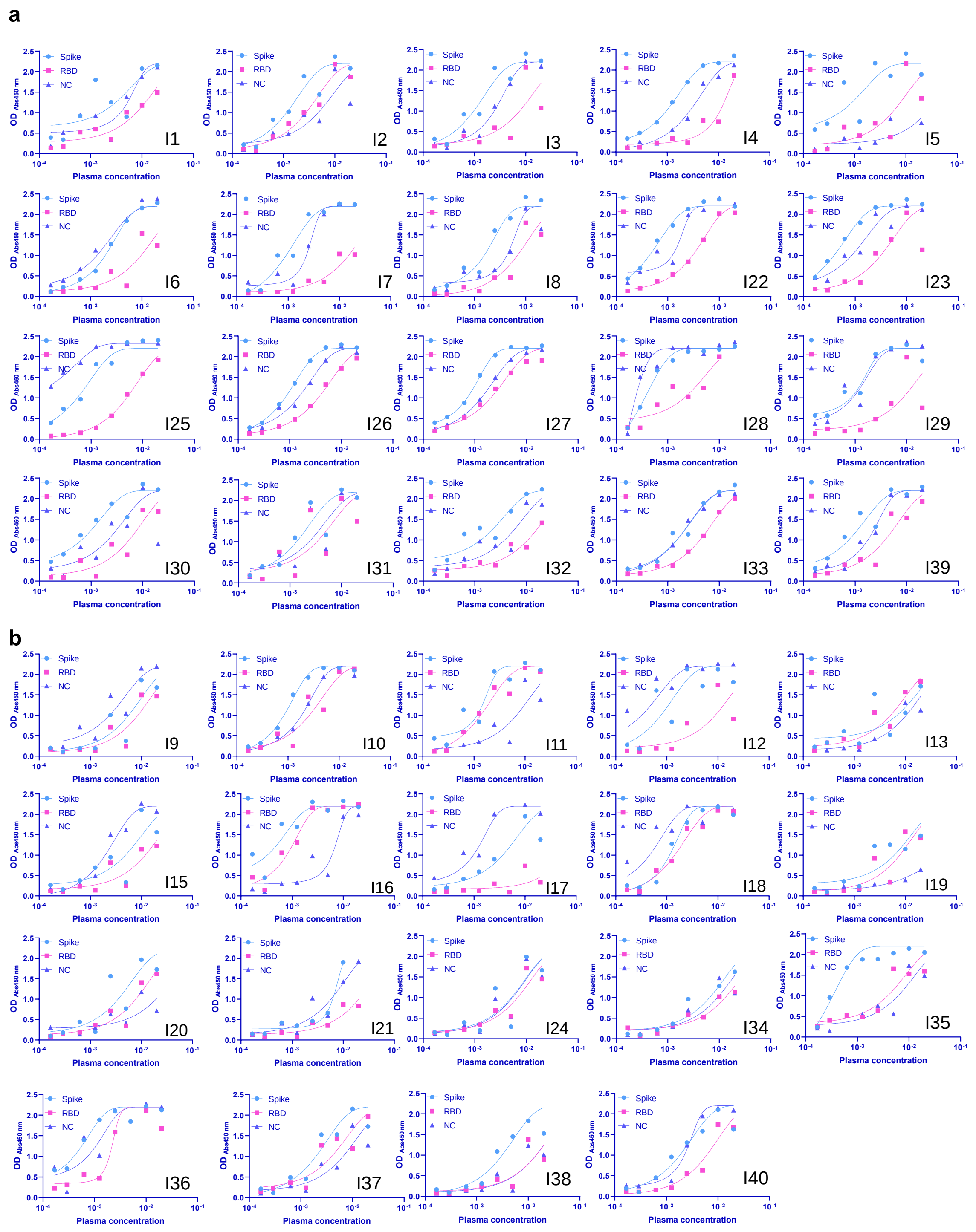


**Figure S1**: ELISA data for the different donors[1]. In light blue, the binding to the spike protein, in pink to the RBD and in blue triangles to NC is shown. (**a**) Convalescent individuals, (**b**) Healthy donors.

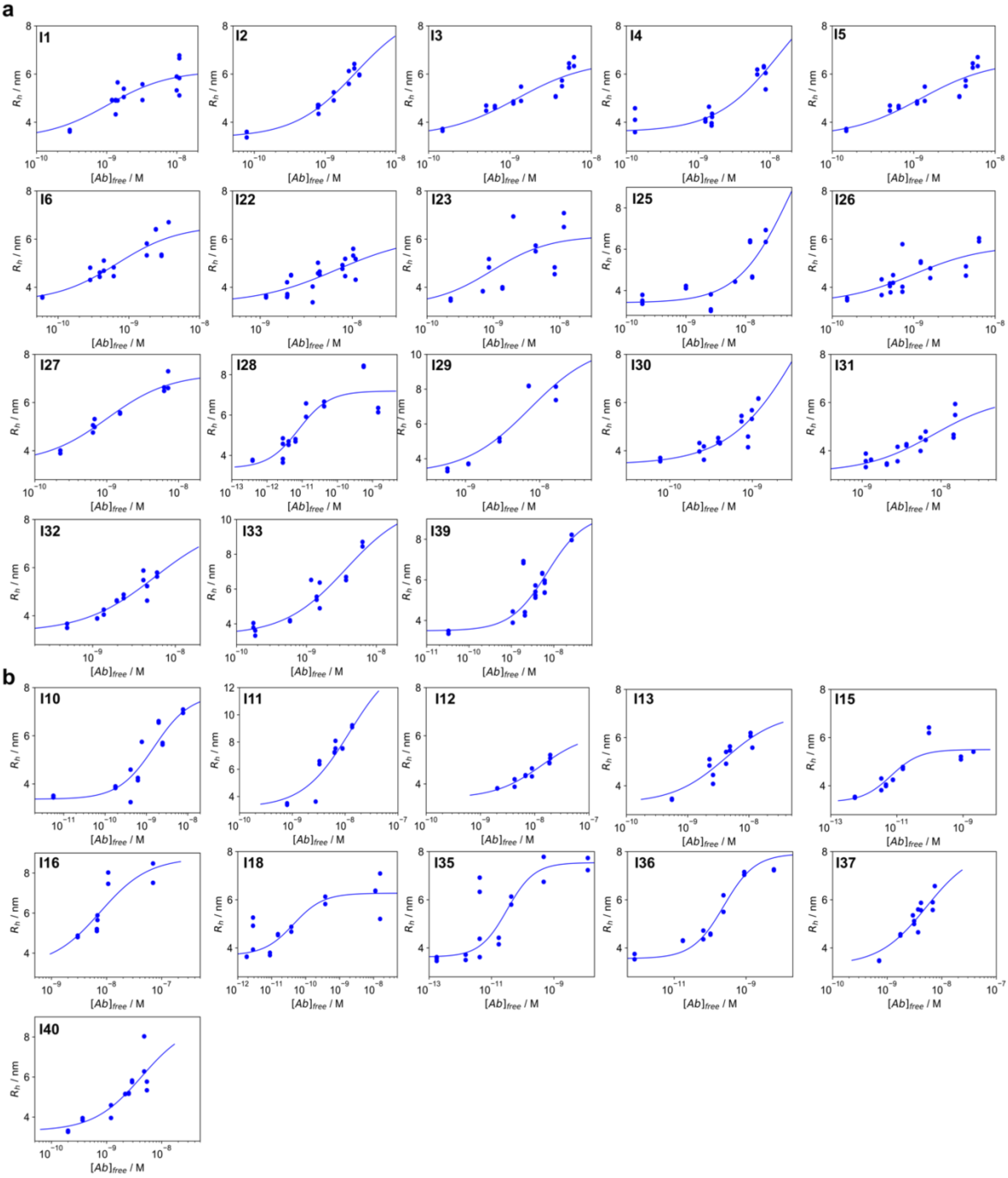


**Figure S2**: Binding curves for all non-hospitalised patients in Fig. 3b. (**a**) Convalescent Donors and (**b**) Healthy Donors.

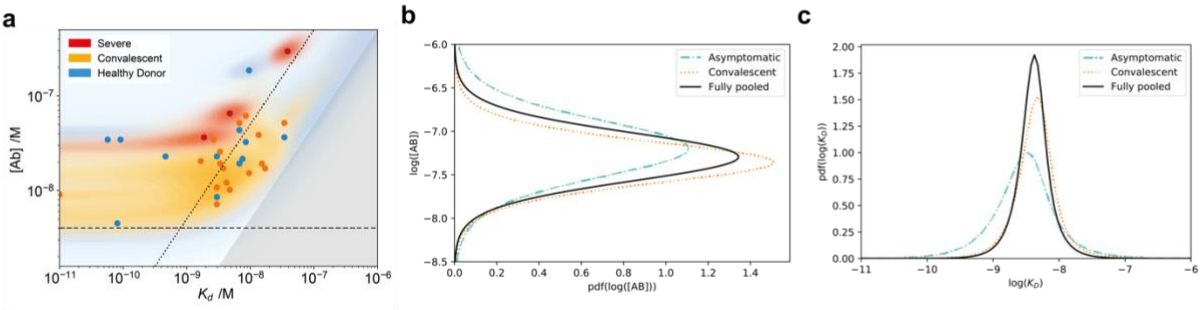


**Figure S3:** (**a**) Probability distributions of dissociation constants, $K_d$, and antibody concentrations in Figure 2B (assuming two RBD binding sites per antibody). Healthy donors (blue), PCR confirmed convalescent (orange), and severely afflicted (red) patients. Points correspond to the maximum probability values in the two-dimensional probability distribution, and coloured regions to the probability density. We do not observe significant difference in either $K_d$ or concentration between different symptom severities. (**b-c**) To address the question whether either log($K_d$) and/or log([$AB$]) differ significantly between asymptomatic and convalescent patients, we analysed the likelihoods from (a) with a partially pooled (grouped by symptoms) and fully pooled hierarchical model[2], as described in methods. (**b**) displays the posterior distribution for $\theta$ = log([$AB$]) and (**c**) for $\theta$ = log($K_d$). The observation that the posterior distributions for asymptomatic (dashed) and convalescent (dotted) patients in the partially pooled model are largely overlapping with one another and the posterior distribution for the fully pooled model (solid) suggests that there are no significant differences between log($K_d$) and log([$AB$]) based on the symptoms experienced.

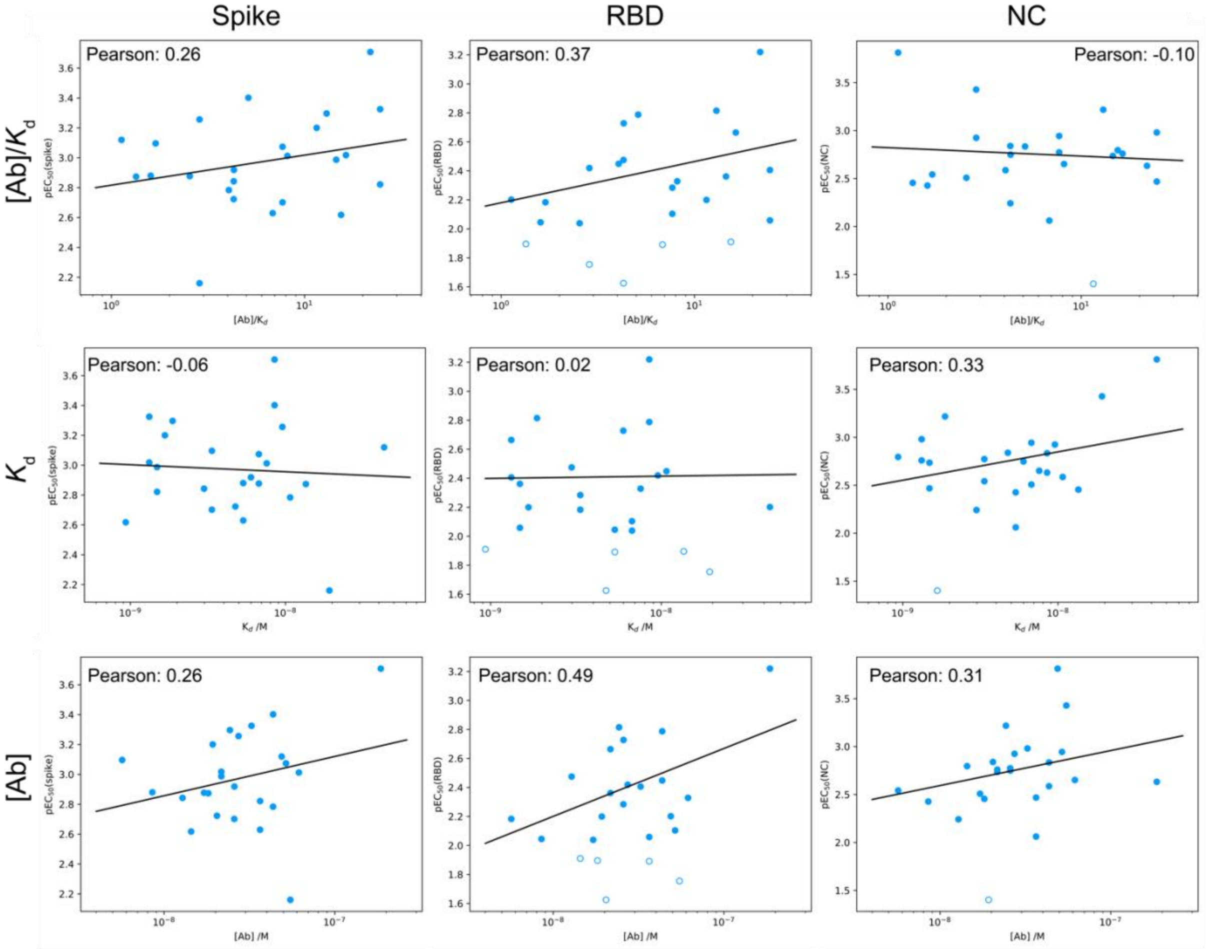


**Figure S4:** Comparison between ELISA (spike: left column, RBD: middle column, and nucleocapsid: right column antigens) and MAAP results for RBD binding. Plots of the pEC$_{50}$ value are shown in comparison to the MAAP-determined ratio of antibody concentration to $K_d$ (top row), $K_d$ (middle row), and antibody concentration (bottom row). Only samples which yielded a peaked posterior distribution in both $K_d$ and antibody concentration are shown, of which only those with pEC$_{50}$ > 2 (filled) were used for the linear regression. Remaining data are shown as open circles. Pearson correlation coefficients are given for each plot.

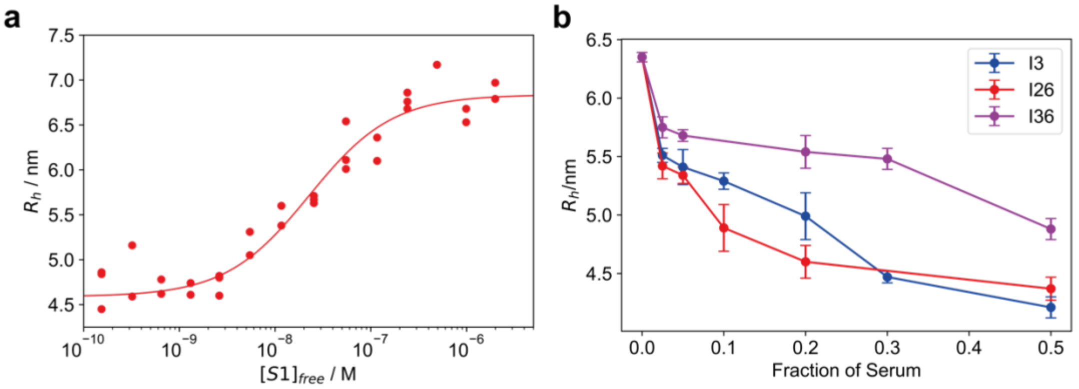


**Figure S5:** ACE2 competition, (**a**) Binding curve of 10 nM ACE2 against fluorescently labelled S1 protein, showing a $K_d$ of 18 [11, 29] nM. Triplicates are shown as individual points. (**b**) Dilution series, showing concentration dependent decrease of hydrodynamic radii in three COVID-19 infected individuals in the competition assay outlined in Figure 3. Error bars are the standard deviation of triplicate measurements.

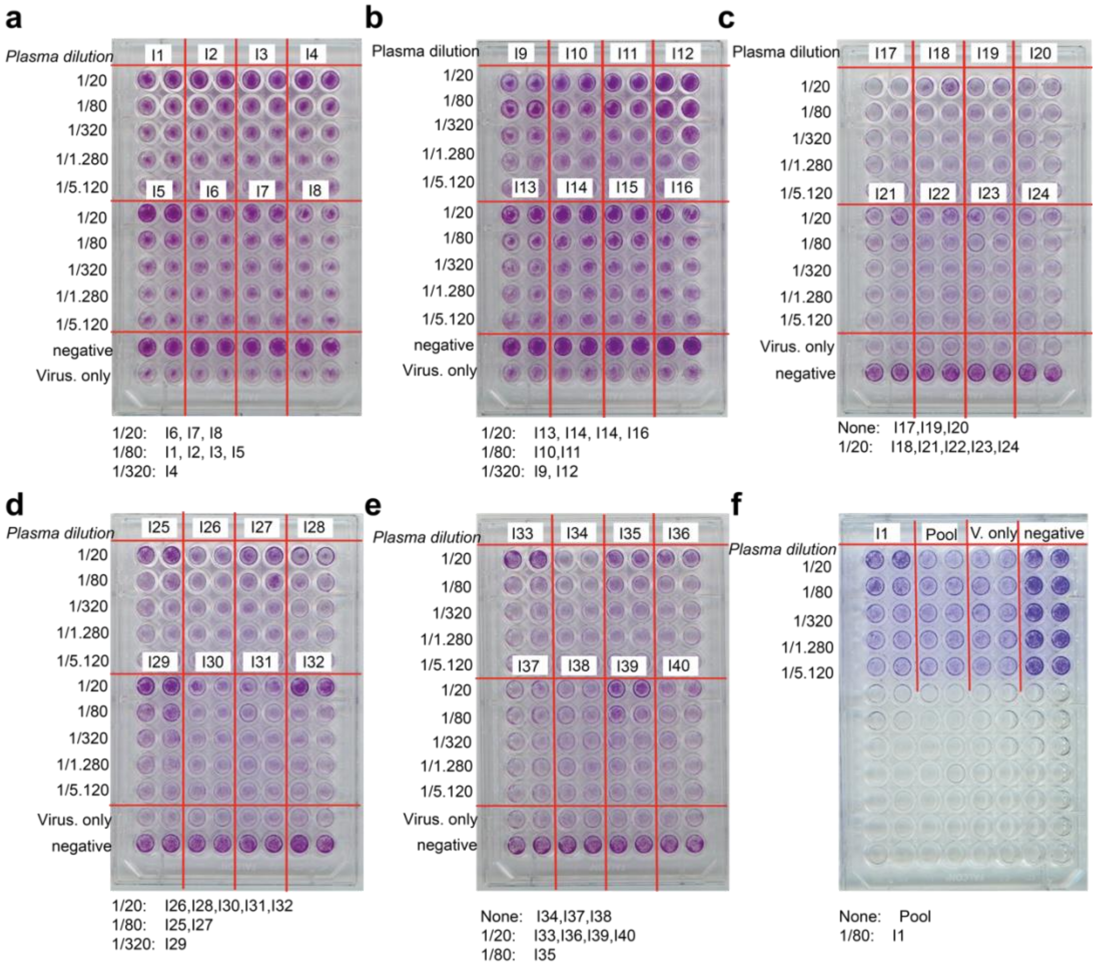


**Figure S6:** Cytopathic effect-based neutralisation assay. Overview over all plates with the respective interpretation of the critical titre below the images.

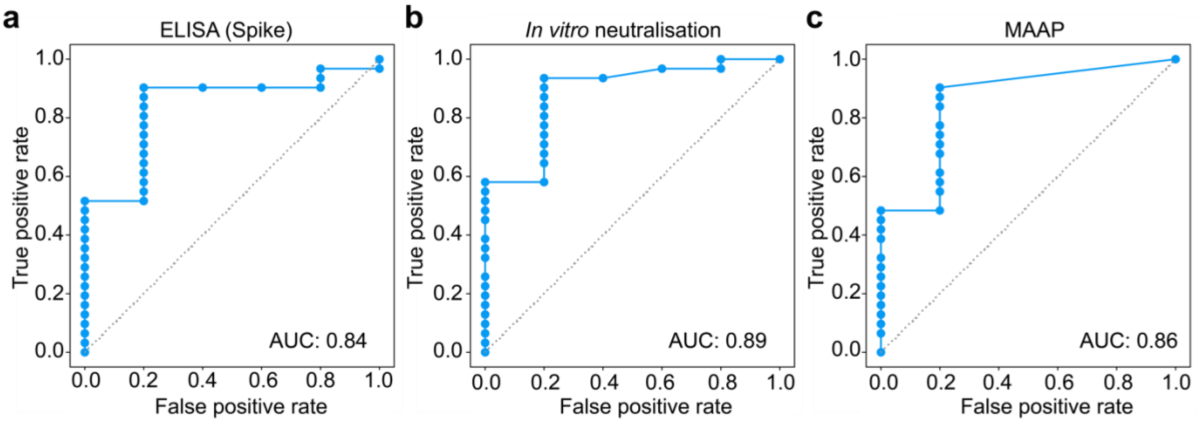


**Figure S7:** Receiver operating characteristic (ROC) curves demonstrating the ability of the ELISA $EC_{50}$(Spike) (a), *in vitro* ACE2 competition assay (b), and MAAP assay (c) to predict the ability of the serum to neutralise in the cytopathic-effect based neutralisation assay, regardless of titre.

**Table S1**. Demographic characterisation of the individuals used in our study.

| **Cohort** | **Total number** | **Number male** | **Number female** | **Number undefined** | **Median age (IQR) - years** |
|---|---|---|---|---|---|
| Convalescent individuals | 19 | 19 | 0 | 0 | 27 (22-49) |
| Healthy blood donors | 20 | 8 | 10 | 2 | 31 (24-45) |
| Hospital patients | 3 | 2 | 1 | 0 | 67 (65-74) |

**Table S2**: Comparison of outcome on ACE2 competition and cytopathic-effect based assay, for all samples which were tested in the ACE2 competition, cytopathic-effect based assay, and had low enough background autofluorescence to be assessed by MAAP. Positive means that the radius has decreased compared to the ACE2-S1 complex. This is the aggregate statistics of the results presented in Fig. S7. A positive result in the ACE2 competition assay is here defined as yielding an effective $R_h$ value smaller than 5.6 nm.

| | | Cytopathic-effect based neutralisation assay | | |
|---|---|---|---|---|
| | | Positive | Negative | Total |
| ACE2 competition | Positive | 29 | 1 | 30 |
| | Negative | 2 | 4 | 6 |
| | Total | 31 | 5 | 36 |

**Table S3**: Comparison of outcome on ACE2 competition and cytopathic-effect based neutralisation assay. Here, we show the hydrodynamic radius based on the ACE2 competition assay as well as the titre of the neutralisation assay. Sera which did not neutralise in the cell plaque assay at any of the titres tested are indicated by 'none', whereas those that were not tested are indicated by 'N/A'.

| Patient ID | ACE2 competition assay $R_h$ /nm | Critical titre for the cytopathic-effect based neutralisation assay (fraction) |
|---|---|---|
| 1 | 4.83 | 0.0125 |
| 2 | 4.98 | 0.0125 |
| 3 | 4.86 | 0.0125 |
| 4 | 5.36 | 0.003125 |
| 5 | 4.84 | 0.0125 |
| 6 | 4.79 | 0.05 |
| 7 | 4.82 | 0.05 |
| 8 | 4.62 | 0.05 |
| 9 | N/A | 0.003125 |
| 10 | 5.14 | 0.0125 |
| 11 | 5.11 | 0.0125 |
| 12 | 4.98 | 0.003125 |
| 13 | 5.18 | 0.05 |
| 14 | NA | 0.05 |
| 15 | 5.06 | 0.05 |
| 16 | 5.25 | 0.05 |
| 17 | 5.76 | none |
| 18 | 5.16 | 0.05 |
| 19 | 5.91 | none |
| 20 | 5.64 | none |
| 21 | 5.67 | 0.05 |
| 22 | 4.74 | 0.05 |
| 23 | 5.14 | 0.05 |
| 24 | N/A | 0.05 |
| 25 | 5.79 | 0.0125 |
| 26 | 4.84 | 0.05 |
| 27 | 5.41 | 0.0125 |
| 28 | 5.07 | 0.05 |
| 29 | 4.77 | 0.003125 |
| 30 | 5.35 | 0.05 |
| 31 | 5.26 | 0.05 |
| 32 | 4.81 | 0.05 |
| 33 | 4.72 | 0.05 |
| 34 | 5.67 | none |
| 35 | 4.96 | 0.0125 |
| 36 | 5.51 | 0.05 |
| 37 | 5.12 | none |
| 38 | N/A | none |
| 39 | 5.30 | 0.0125 |
| 40 | 5.08 | 0.05 |
